# UK Health Care Workers’ Experiences of Major System Change in Elective Surgery during the COVID-19 Pandemic: Reflections on Rapid Service Adaptation

**DOI:** 10.1101/2021.04.14.21255415

**Authors:** Georgina Singleton, Anna Dowrick, Louisa Manby, Harrison Fillmore, Aron Syverson, Sasha Lewis-Jackson, Inayah Uddin, Kirsi Sumray, Elysse Bautista-González, Ginger Johnson, Cecilia Vindrola-Padros

**Author notes:** Joint first authors (Georgina Singleton, Anna Dowrick).

## Abstract

**Background:** The COVID-19 pandemic disrupted the delivery of elective surgery in the UK. The majority of planned surgery was cancelled or postponed in March 2020 for the duration of the first wave of the pandemic. We investigated the experiences of staff responsible for delivering rapid changes to surgical services during the first wave of the pandemic in the UK, with the aim of developing lessons for future major systems change.

**Methods:** Using a rapid qualitative study design, we conducted 25 interviews with frontline surgical staff during the first wave of the pandemic. We also carried out a policy review of the guidance developed for those delivering surgical services in pandemic conditions. We used framework analysis to organise and interpret findings.

**Results:** Staff discussed positive and negative experiences of rapid service organisation. Clinician-led decision making, the flexibility of individual staff and teams, and the opportunity to innovate service design were all seen as positive contributors to success in service adaptation. The negative aspects of rapid change were inconsistent guidance from national government and medical bodies, top-down decisions about when to cancel and restart surgery, the challenges of delivering emergency surgical care safely and the complexity of prioritising surgical cases when services re-started.

**Conclusion:** Success in the rapid reorganisation of elective surgical services can be attributed to the flexibility and adaptability of staff. However, there was an absence of involvement of staff in wider system-level pandemic decision-making and competing guidance from national bodies. Involving staff in decisions about the organisation and delivery of major systems change is essential for the sustainability of change processes.

## Introduction

The COVID-19 pandemic has placed unprecedented strain on healthcare systems around the world. In the UK, rapid reorganisation of care delivery was required due to the National Health Service’s (NHS) inability to cope with patient demand in the context of limited critical care capacity. One of the strategies used to increase capacity was the cancellation and postponement of elective surgery. This enabled a number of other changes. Hospitals were able to use operating theatres and recovery areas as “surge areas” where intensive care units (ICUs) could expand, theatre staff were liberated for redeployment and anaesthetic machines were made available for ventilation. Moreover, the flow of patients who did not require urgent care in hospitals was limited, thereby reducing the incidence of intra-hospital infection.

The reorganisation of care delivery at a national scale and within short timeframes provides an opportunity to examine the factors that can act as barriers and enablers of major system change (MSC) in healthcare systems. MSC involves the planning and implementation of new pathways of care^1^ and shifts in ways of working across multiple healthcare organisations^2^. Existing research in this field has emphasised that a significant contributor to MSC success is the ability of actors involved in implementing change to make adaptations in line with the opportunities and constraints of their local context^3^.

While there has been substantial description of the overall changes to elective surgery that took place during the pandemic^4–6^, there is less understanding of healthcare workers’ (HCWs) experiences of implementing these service adaptations. Understanding how HCWs navigated and enacted challenging decisions regarding the redesign of elective surgical services during the pandemic will provide lessons for the planning for future MSC, particularly during pandemic conditions.

In this paper, we analyse the experiences of staff involved in adapting the delivery of elective surgery in three UK hospitals during the first wave of the COVID-19 pandemic. In the first part of the paper, we report on the policy context of surgical care during the pandemic. In the second, we provide insights into the positive and negative experiences of reorganisation as perceived by those involved in service delivery during this period of rapid change.

## Background

### Preparedness for delivering elective surgery during a pandemic

Delivering an effective surgical service in the UK NHS was challenging even before the pandemic. A decade of austerity in national spending^7,8^ had led to the lengthening of waiting times for elective surgery^9^ and growing incidence of cancellation^10^. The “UK *Influenza Pandemic Preparedness Strategy”*^11^ recognised that, in the case of a severe pandemic, routine elective surgery would be seriously affected. This and other similar policy documents^12,13^ put an emphasis on hospitals making local decisions about how to reorganise services in the event of a pandemic. A pandemic preparedness simulation run in 2016 identified the need to model the full impact of service closures to inform decision-making as well as the need to develop plans to support the communication of such decisions^14^.

These strategies were put to the test by an unexpectedly severe epidemic of seasonal flu in the winter of 2017/2018. In order to cope with this, all elective surgeries were cancelled, which included all planned, routine non-cancer surgeries from December 2017 to January 2018, leading to a large backlog of surgical procedures^7^. The advice provided by the NHS stated that each hospital could decide exactly how to manage the reorganisation of services at this time^15^. In August 2018, this backlog led the NHS to develop an urgent plan to redirect “significantly more” patients to private healthcare providers for their routine procedures^16^. While this preparation and experience gave some insight into pandemic conditions, further detailed preparations for a pandemic response were not undertaken in UK hospitals.

### The COVID-19 pandemic and UK elective surgery policy change

With the onset of the COVID-19 pandemic, NHS England requested that staff suspend non-urgent elective operations in preparation for the predicted rise in demand for beds, resources and staff availability^17^. The Royal College of Surgeons (RCS) published guidance on the 20^th^ March 2020, which recognised the need for the surgical workforce to adapt during the COVID-19 pandemic^18^.

This approach for managing hospital capacity was similar to that taken in the US and Italy, where hospitals reduced operation room schedules and removed patients planned for non-essential procedures from operating lists^19,20^. Other countries, such as South Korea and Singapore^21^, continued with elective work during the pandemic by adding new control and monitoring measures, such as screening and testing patients before admission and reorganising surgical work into ‘hot teams’ managing acute surgical admissions and ‘cold teams’ continuing with elective surgical work.

The RCS recognised the difficult decisions the surgical workforce had to make with regards to prioritising surgical procedures and released further guidance on the 11th April 2020 to aid with the decision-making process ^4^. They allocated priority levels to various forms of surgical procedures to help guide the allocation of finite numbers of resources and staff. It was advised that patients needing urgent surgery (such as emergency admissions or cancer treatment), patients who had previously had their procedure delayed, and children should be prioritised ^4,17^.

In an attempt to increase labour, resources and facilities to allow the continuation of essential services such as cancer and clinically urgent surgeries, the NHS gained extra capacity from independent, private hospital providers. Private-to-public hospital conversion for surgical care was operational in the UK from the 23rd March 2020. This enabled the NHS to transform independent hospitals into ‘COVID-19 light sites’^22^. These sites were specific units or hospitals that provided elective surgical care for non-COVID-19 patients.

### Impact of COVID-19 on elective surgery

Estimates suggest that just over 43,300 surgeries were cancelled each week during the 12-week period in which elective surgical procedures were suspended during the first wave of the pandemic in the UK^21^. With the mounting backlog of procedures that this created, the UK set out to restart elective surgeries on 18 June 2020^23^. Public Health England (PHE) published infection and prevention control guidance which outlined that patients undergoing planned/elective surgical procedures should follow a low-risk COVID-19 pathway^24^. This meant that patients would need to test and isolate prior to their surgery as well as undergo a clinical assessment of symptoms before their procedure. In addition, emergency rotas were developed and new roles were advertised to reduce the burden of current staff workload and maintain the quality of patient care^25^.

As elective surgeries restarted in the UK, NHS England set targets that aimed to achieve 80% of the previous year’s surgical activity by the end of September and 90% by the end of October 2020^5^. However, even with established COVID-safe pathways and outsourcing to COVID-light sites, a report published on 6 October by the RCS indicated that surgeons were struggling to reach these targets^26^. A lack of theatre capacity and sub-optimal levels of staff were major contributing factors. Later in October, several Trusts across the UK reviewed their position and made the decision to re-suspend non-urgent elective procedures in order to cope with the increasing number of COVID patients during the UK’s second wave of the pandemic^27^. The policy timeline of decisions about suspending and re-starting surgery is summarised in figure 1.

**Figure 1:**
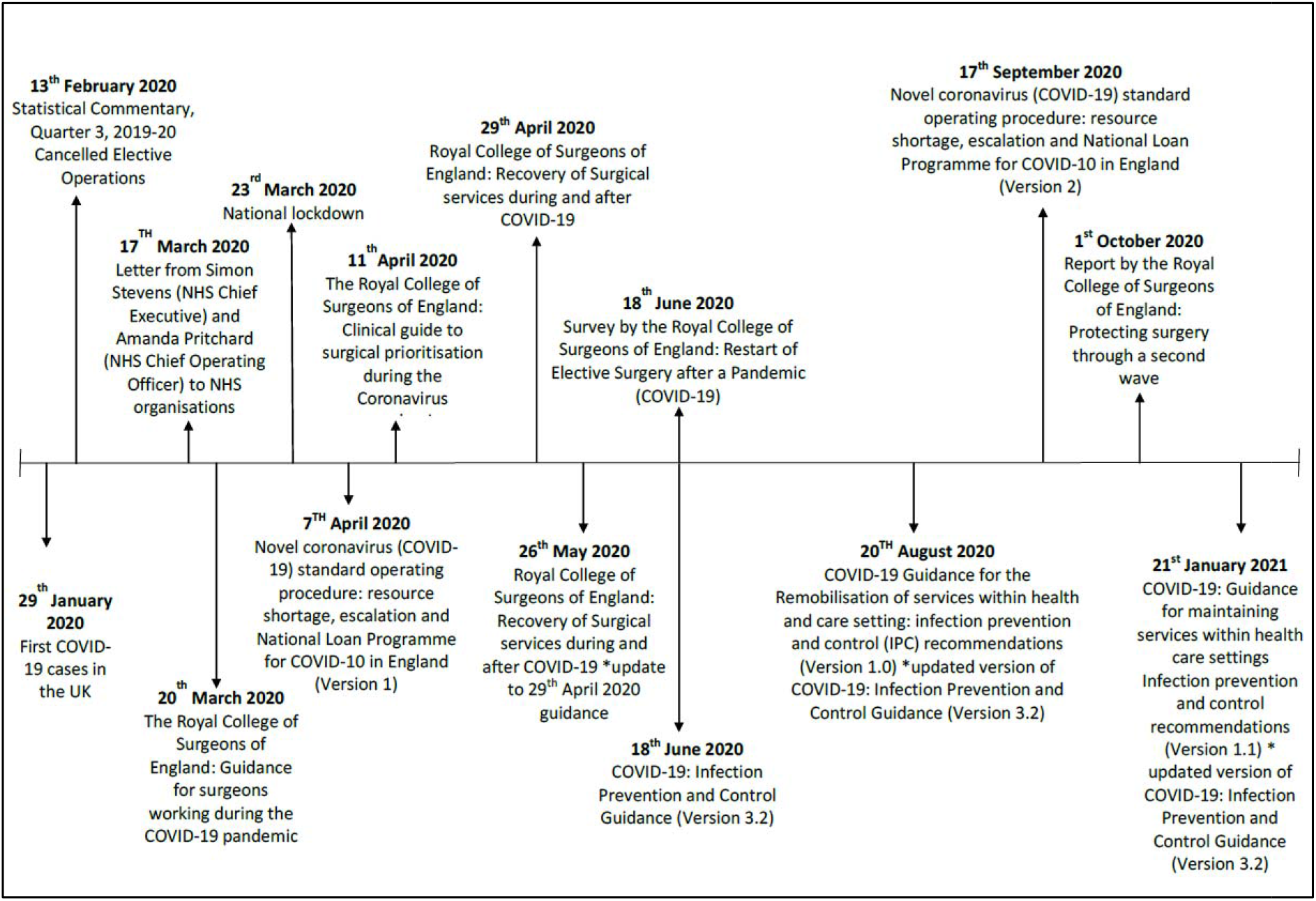
Timeline of events impacting the delivery of elective surgeries in the UK.

**Figure 2:**
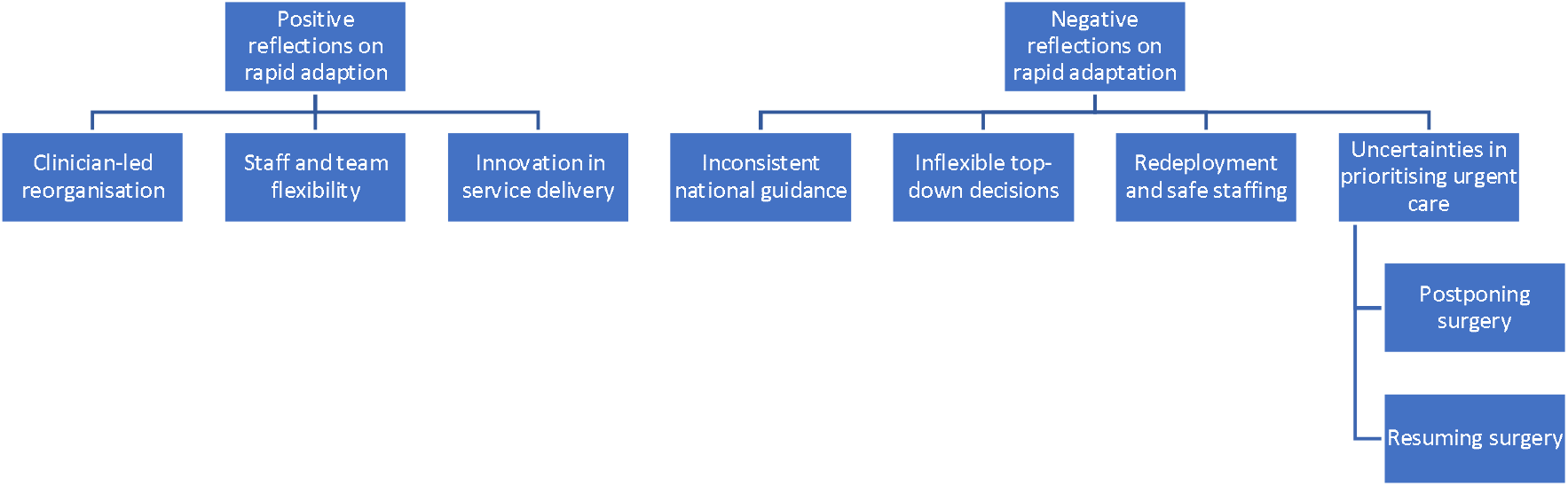
Summary of themes

### Reorganisation of elective surgery as major systems change

We frame the rapid change in the delivery of elective surgery in the UK during the first wave of the pandemic as an example of major systems change (MSC). MSC involves introducing specific, targeted changes to the organisation of work within systems which affect a wide number of staff, often framed around the concept of re-designing ‘pathways’ of care for patients^2^. Jones et al^28^ define these processes as:

*“Policies, strategies or interventions that aim to transform the way multiple care services are coordinated at the inter- and intra-organisational level to address a single service area (e*.*g. stroke) or integrated service domain (e*.*g. primary care)*.*”*

The MSC field to date has focused on examples of planned system changes, such as centralising service provision^1,29,30^, integrating different forms of public service^31^, and introducing new technologies into health systems^32^. The case study of changes to elective surgery provision during the pandemic enables investigation of the enablers and consequences of unanticipated, rapid change.

While there are a number of characteristics identified as enablers of MSC - unified leadership, locally driven decisions, rapid monitoring and feedback to enable adaptation, and engagement of staff and patients^33^ – it is how these characteristics dynamically manifest in a specific local context that is of particular importance to the success of change processes^34^. Involvement of those who will be responsible for implementing MSC in decisions about adapting local practices has been identified as crucial to successful implementation^35,36^, as reorganising service delivery is ultimately achieved by multiple groups of staff collectively adapting their locally established working practices and relationships^28^. With regard to changes in the provision of surgical services, there has to date been limited investigation of the experiences of staff who implemented these changes. There is growing emphasis on the importance of learning from the adaptations of staff involved in delivering new practices^1,3,37^, which we seek to build on and contribute to through this research.

In this paper, we will contribute to MSC scholarship through an examination of UK HCWs’ experiences of reorganising and adapting elective surgical services in response to the COVID-19 pandemic. We focus our analysis on staff’s reflections about the positive and negative aspects of rapid service organisation, drawing attention to the perceived barriers and enablers of adapting to pandemic conditions. We provide practical lessons for policy-makers and insights about the challenges of implementing MSC in pandemic conditions.

## Methods

Adopting a theoretical perspective that MSC is enacted through social practice^38^, we aimed to develop insights from the perspective of HCWs involved in enacting changes in surgical services during the pandemic. We argue that understanding the enablers of rapid change and reorganisation during the pandemic from the perspective of those delivering it contributes broader insights for how policy can guide the effective and safe delivery of elective surgery throughout the duration of the pandemic and beyond.

This study was part of a larger, ongoing programme of research investigating the perceptions and experiences of HCWs during the COVID-19 pandemic. The wider study utilises a rapid appraisal methodology integrating data from three research streams: telephone interviews with frontline HCWs, UK policy review and media analysis^39^. For this paper, we conducted a policy review and drew on in-depth, semi-structured interviews with frontline staff.

### Policy review

The aim of the healthcare policy review was to understand how surgical service delivery has been reorganised as a result of the COVID-19 pandemic in the UK, exploring the advice given during the first wave of the pandemic and subsequent reflections as the UK entered a second wave. We followed the framework set out by Tricco et al^40^ for rapid evidence synthesis. We searched for government policies on public NHS databases using the following search strategy:

Search strategy for UK policy review:

Search terms: COVID-19 OR coronavirus OR corona AND surgery Inclusion criteria:

1. Published from 1 December 2019 to 1 November 2020;
2. Aimed at surgical healthcare delivery
3. Related to the COVID-19 pandemic.

CVP selected the policies that met these criteria. SLJ, GS, LM and IU reviewed the policies and extracted data regarding the type of policy, professional group it was aimed at, the type of changes in service delivery it proposed and the duration of these changes. Data were cross-checked across reviewers. The policy review informed the background section of this paper.

### In-depth interviews

The aim of the in-depth interviews was to capture the experiences and perceptions of frontline surgical staff in relation to the impact of COVID-19 on the delivery of surgical procedures. Semi-structured interviews were carried out from 28 March 2020 to 26 June 2020 using a semi-structured topic guide (see Appendix A) which focused on the impact of COVID-19 on elective and emergency surgery delivery, the decision to cancel elective surgeries and the preparedness strategies in place to guide this process as well as the concerns or fears related to the restarting of elective surgeries.

Interviews were conducted via telephone with a purposive sample of 25 frontline staff across 3 London trusts which were involved in the wider research programme^41^. Potential participants were identified by local hospital leads. A sampling framework was developed in order to guide recruitment which included different professional groups, levels of seniority and gender. The final sample included anaesthetists (11), surgeons (5), nurses (4), surgical trainees (2), surgical assistants (2) and a service manager (1). Over half of the sample was female, with the majority of participants in middle or senior management positions. Years in practice ranged from less than a year to thirty-two years in service.

These individuals were provided with a participant information sheet and were asked whether they were interested in being contacted by a member of the research team. Where individuals expressed an interest in participating, they were contacted by a researcher asking if they had any questions about the study. When HCWs agreed to take part in the study, they were asked to sign a consent form. Participants were reminded that their participation was voluntary and that they could withdraw from the study at any time and maintain anonymity. Interviews were audio-recorded and transcribed using an authorised transcription service.

Interview notes were imported into rapid assessment procedures (RAP) sheets, which were used to synthesise findings on an ongoing basis^41^. Researchers (KS, AD, AS, HF, LM, EBG) collectively read transcripts and developed an analytical coding framework based on a preliminary scan of the data using a framework analysis approach ^42^. The coding framework focused on identifying HCW’s positive and negative reflections on MSC during the pandemic. These codes were inputted into a Microsoft Excel matrix, with the emerging codes in the columns and interviews entered as individual cases in the rows. The framework was refined during team discussions and all researchers were asked to apply the same framework across their assigned interview transcripts. KS, AD and GS cross-checked the data during the coding process to ensure consistency across researchers.

The study was reviewed and approved by the Health Research Authority (HRA) (IRAS: 282069) and the R&D offices of the hospitals where the study took place.

## Results

### Positive reflections on rapid adaptation

#### Clinician-led reorganisation

Participants in this study attributed the success of the rapid reorganisation of services to changes being clinician-led. While the decision to cancel or delay the majority of elective surgeries ‘*came from the top’* (COV74: Anaesthetist), the work of deciding which surgeries to cancel, how to modify care pathways to enable emergency surgeries to continue, and how to re-purpose surgical wards into makeshift intensive care units (ICUs) was done by clinicians. Staff reflected that *‘it was a little bit chaotic at times but if you look back, it actually worked quite well’* (COV38: Surgeon).

Staff were able to adapt service delivery according to their immediate clinical context. Moreover, usual management-led organisational bureaucracy was to a large extent circumvented. This was also reflected in the liberal allocation of resources, including improved provision of parking and accommodation: ‘*there is free parking now and multistorey places to park and also there hotels in place for you to stay’* (COV26: Healthcare Assistant). Changes that would have *‘in normal times taken decades probably to get into place’* (COV8: Anaesthetist) were achieved in the space of weeks.

Decisions about how to effectively suspend surgeries and reallocate resources were made at trust and regional levels, as opposed to nationally. This was facilitated by ‘*heavy clinical leadership during the pandemic, with teams collaborating together across sites, across hospitals, across specialties’* (COV97: Surgeon). This approach enabled consideration of local context, in relation to the surgical infrastructure available and the impact of COVID-19 in a given area at a particular time. For example, some hospitals involved in this study collaborated with other regional hospitals to transform some services into cancer hubs which maintained urgent cancer care.

> *Surrounding hospitals would redirect their patients to us, because we were supposed to be COVID free, and we somehow became a cancer hub basically for the north of the city*.
>
> COV94: Anaesthetist

A major contributor to rapid adaptation was the ability of staff to collectively draw on their clinical and professional experience to redesign service delivery, collaborating across departments and hospitals to reallocate resources.

#### Staff and team flexibility

Teams displayed agility in adapting to rapid restructuring, with roles, teams and rotas all regularly changing. Some surgical staff were redeployed to support ICU services while others worked to maintain the provision of emergency care, with regular adjustments to reflect the loss of staff due to illness or requirements for self-isolation.

> *We have people who worked Week A and Week B essentially so that if anyone was ill it keeps some of the workforce separate, so we would always have a core workforce who weren’t ill*.
>
> COV45: Surgeon

Senior staff in particular were more active in supporting colleagues, with more consultants present overnight to oversee services - ‘*we used to have no consultant anaesthetist resident overnight, we now have 10 resident in the hospital doing various things’* (COV24: Anaesthetist). In some instances, this meant taking responsibility away from junior team members in order to increase efficiency, for example suspending training for junior surgical staff.

> *We decided that we would be operating ourselves, between consultants, in order to minimise the risk of complications and in order to speed up the operations*.
>
> COV97: Surgeon

Delivering change quickly was facilitated by improved communication within and between teams - *‘there was a chain of communication from, from the top, several times a day, every day’* (COV94: Anaesthetist). Rapidly understanding the needs of different teams enabled staff to identify where there were gaps in training or equipment and to reallocate resources accordingly.

> *There was lots of efforts in grassroots. I was involved in training of airways, in the end I just did it myself because we couldn’t wait any longer, for the collective benefit of our colleagues and patients. Management have been very supportive but it is clinicians who have been coming knocking on the door saying we need to prepare and perform these trainings*.
>
> COV19: Anaesthetist

Identifying and responding to local needs regarding the provision of resources, support and training meant that teams could rapidly restructure themselves to adapt to pandemic conditions.

#### Innovation in service delivery

Despite the challenges of reorganising services, staff felt the way hospitals had been able to react was ‘*very interesting and exciting’* (COV37: Service Manager). A great deal of improvements were thought to stem from the redesign and adaptation of services carried out during the peak of the COVID-19 pandemic.

> *Even after the pandemic we definitely want to keep the good things, all the changes we’ve implemented, because we’ve seen tangible results in our everyday practice*…*in terms of time-saving, resource-saving, increasing efficiency, more appropriate managing of resources, managing of staff, all that*.
>
> COV97: Service manager

This was felt particularly in relation to the flexibility and collaboration between teams and sites, as discussed above, but also with regard to new approaches to caring for patients remotely. Efforts to reduce flow of patients through hospitals meant that staff introduced remote consulting to deliver pre-operative care.

> *We weren’t doing many telephone consultations before but I think we’ve realised that there are certainly some patients in the future who will be suitable for telephone follow-ups after this which we would have taken a while to realise otherwise*.
>
> COV45: Surgeon

While there were frustrating experiences, such as problems with technology, several HCWs stated that the push into digitalisation during COVID-19 may be beneficial to the wider acceptance of remote care across the perioperative pathway:

> *It’s been a good opportunity to streamline that, because there’s a lot more remotely and in a streamlined process, that we can do it all in one or one or if not, no hospital visits, and do a lot of online, and a lot of it remotely. So hopefully it’s something we’ll be able to take forward um, once it’s all over. Not do it to the same level we’re doing it now, but certainly use some of those aspects that keep the health service more efficient*.
>
> COV10: Surgeon

Learning from this experience was also seen as beneficial for preparations for a second peak or a future pandemic. For ongoing uptake of new approaches to be successful, staff suggested that training in innovative practices should continue even when COVID-19 cases are few, informed not only by the UK response but also by insights from other countries.

### Negative reflections on rapid adaptation

#### Inconsistent national guidance

Throughout the COVID-19 pandemic, professional societies and national organisations in the UK offered guidance about delivering elective procedures. This guidance was not felt to be sufficiently clear about how to maintain safety during the surgeries that did proceed.

> *We were getting different information every day from different sources, from Europe, within maternity, within the Trust, from PHE, and I think everything felt very, very different…I found that a bit stressful, this conflicting advice and nobody being quite sure what’s the right thing to do. Do we break the Trust rules and do what we think is right ourselves?*
>
> COV45: Surgeon

Inconsistent guidance made staff feel in conflict with national bodies. At times they felt they had to fight to protect themselves, for instance in having to negotiate access to PPE for surgical procedures they deemed to be risky but were not recognised in formal guidance as such. Beyond safety as teams, some participants also felt that they were left to risk-assess their individual risk of COVID-19 exposure.

> *I mean that just seems absolutely extraordinary and we put the onus on individuals to say ‘I don’t think I’m safe to work in COVID areas’, so it was almost like saying, ‘it’s your problem’. We weren’t building it into the system to protect our staff*.
>
> COV95: Anaesthetist

While staff attributed success in service reorganisation to clinician-led decisions about adapting service delivery, they sought more guidance about delivering surgery safely.

> *There should be clear guidance from NHS and Government running down to Trust level rather than having to wait for clinicians to change things themselves*.
>
> COV19: Anaesthetist

Staff argued that local decisions would have been easier within the context of consistent national guidance providing the underlying principles of safe care during the pandemic.

#### Inflexible ‘top-down’ decisions

The overall decision to postpone non-urgent surgery was considered by the majority of participants to be a ‘top-down’ decision, with little involvement from frontline clinicians.

> *We didn’t have much of a say when it came to what was going to get done, whether it was going to get stopped, and when it was going to restart…, we didn’t really have that input, or that platform to voice our concerns. It was just ‘this is what we’re doing, you guys have to do it’*.
>
> COV74: Anaesthetist

While the decision to change the model of elective surgery delivery during the pandemic was not contentious among participants in this study, there was ambivalence about the exact way in which services were reorganised. Some staff felt that the decision to stop all elective surgery was ‘*the right one’* (COV85: Surgeon) because of the uncertainty around the risks of surgery at the outset of the pandemic. Equally, other participants reflected that there may have been other approaches for managing risk while continuing to provide surgical services.

> *We may not have shut down the whole service in the way that we did. I think we could have had a more strategic plan*.
>
> COV95: Anaesthetist

This uncertainty was motivated by concern about the future consequences. Some feared that the NHS was ‘*stretching a different problem further down the road’* (COV38: Surgeon), with a growing backlog of support needed for patients whose procedures were postponed. Some of the challenge related to the speed at which the decisions were made. Staff noted in particular the delay between awareness of the impact of COVID-19 in Europe and subsequent decisions made about the UK response:

> *We could have had a more strategic plan if, as a group we decided to say ‘we’re going to need to model what’s happened in Italy across London but we’re going to need to keep elective surgery going’, rather than holding off, holding off and then doing a kind of crisis response within a four to six week period*.
>
> COV95: Anaesthetist

HCWs in this study shared a commitment to adapting elective surgery service delivery in light of the pandemic, but displayed considerable doubt in their reflection on both the decision by NHS England and the Royal College of Surgeons to cancel all but essential surgical services and its timing.

#### Redeployment and safe staffing

While staff redeployment was considered key to the pandemic response, it was noted to cause anxiety, confusion and stress. Starting work in ICU without appropriate skills for their role was difficult for redeployed surgical staff. While formal training was made available, there was not always time to undertake it.

> *There’s a lot of anxiety amongst the theatre staff and the operating staff and they’re all feeling a bit overwhelmed by their experiences in the make-shift ITU that we created. They’re all not feeling great*.
>
> COV95: Anaesthetist

Moreover, redeployment was felt to reflect service needs rather than staff skills.

> *They redistributed nurses every day. Respiratory trained nurses which would have a better understanding of respiratory problems than a surgical nurse, but those nurses aren’t being redeployed to ICU. They were not looking at skill sets, treating each nurse as a number rather than looking at the skills of nurses*.
>
> COV22: Ward Sister

The changing composition of teams, with *‘new faces within the department every day’* (COV94: Anaesthetist), also made it difficult to establish working practices as a group. Keeping track of rapidly changing processes was also a challenge: ‘*it was really exhausting just trying to work out what we were doing with different things coming up in different places all the time*’ (COV45: Anaesthetist).

High levels of staff sickness, which were felt to be ‘*decimating our service’* (COV10: Surgeon) resulted in higher intensity working patterns for those who remained at work. This impacted on the perceived quality and efficiency of surgical teamwork.

> *We used to be a really slick oiled team, we’d have everything ready, but you can see they’re all worn out*.
>
> COV95: Anaesthetist

The concern among all participants was how to manage both surgical activities and COVID-19 care, as it was felt that the hospitals did not have enough staff to *‘provide a full set of normal services alongside COVID services’* (COV7: Anaesthetist). Staff felt they were being asked to deliver two parallel health systems without sufficient resources: one for COVID-19 and one for non-COVID-19 care.

#### Uncertainties in prioritising urgent care

##### Postponing surgery

Enacting a reduction in provision of elective surgery involved staff making decisions about which patients’ surgeries would be postponed. While the RCS provided guidance on which categories of surgeries should be prioritised, such as surgeries for adults requiring urgent care, staff had to contextualise these decisions within the lives of individual patients, with staff *‘very anxious about not exposing people to major surgery and COVID at the same time’* (COV95: Anaesthetist). This involved making a judgment about what constituted ‘urgent care’, balancing the impact of delay on patient outcomes against the demands COVID-19 was placing on the healthcare system.

> *My neighbour she was due to have surgery at my hospital next week for a massive prolapse that is painful, that is… it’s not life-threatening but it’s not life if you cannot walk anywhere because your organs are falling out from your holes, you know. So, she cannot have it and we don’t know when she’s going to have it, and this is for so many patients*.
>
> COV11: Surgeon

Similarly, HCWs were aware of the potential risks of continuing with procedures, for patients and for themselves, particularly during aerosol generating surgeries which were ‘*highly contagious’* (COV74: Anaesthetist). For patients who attended in-person appointments, HCWs described higher risk during procedures and higher levels of distress for patients pre- and post-operatively. Some HCWs attributed this to not being allowed someone to accompany them to the hospital and was likely exacerbated by the impersonal nature of being attended to by staff wearing full PPE.

> *Post-operatively then they’re not allowed any visitors and I think that definitely impacts on their mental health*.
>
> COV97: Surgeon

Deciding whose surgeries would continue involved complex judgment of what discomfort could and should be tolerated by patients, as well as a reflection on the risk of proceeding for both patients and staff.

##### Resuming surgery

HCWs restarted elective surgery following the reduction in COVID-19 cases after the first national lockdown. This represented another major change to service delivery, with staff reporting that it was *‘harder to restart elective work than it was to stop and create a new ITU’* (COV95: Anaesthetist).

Healthcare staff were widely concerned about the backlog of surgeries and waiting times for patients following the reintroduction of elective and non-urgent surgical procedures. While staff had been able to categorise cases as ‘urgent’ in the early stages of the pandemic based on immediate threat to life, they faced a significant challenge in deciding where to start among those patients whose surgeries had been postponed: ‘*we don’t really understand how to prioritise’* (COV26: Anaesthetist).

Staff were faced with a number of concurrent challenges. As well as patients whose care had been delayed, other patients needed surgical interventions as a result of their conditions worsening after avoiding hospitals during the pandemic. Decisions about prioritising ‘*who goes on the waiting list first’* (COV45: Surgeon) were taxing. Staff had to weigh up factors that had no clear equivalence, such as whether to prioritise patients based on their risk of further complications, how long they had been waiting, or their current level of pain:

> *So, for example, many patients will be on a list for orthopaedic surgery, so all the patients who … who you know, who have two new hip replacements and knee replacements, things like that, which are debilitating conditions, which aren’t necessarily big and life threatening, aren’t getting done. But yet they’ll still be at home, in their sort of pain or whatever, having to deal with it*.
>
> COV10: Surgeon

Both postponing and restarting surgery involved complex decision-making that left staff unsure about how to ‘*give people waiting for surgery equitable healthcare’* (COV32: Anaesthetist).

As well as trying to fairly balance the needs of patients suffering in different ways, staff also had to balance the scale-up of surgery against the potential of a return to high levels of COVID-19 infections. This involved decisions about ‘*how to do that [reintroduce surgeries] safely and effectively but not get too ahead of the game in case there is a second spike’* (COV32: Anaesthetist).

## Discussion

This study aimed to understand the experiences of staff adapting the delivery of UK surgical services during the first wave of the COVID-19 pandemic, interpreting their experiences as a case study of MSC. The success of service adaptation was attributed to clinician-led reorganisation of care into different hubs of localised service delivery which addressed the emergency surgical needs of patients with and without COVID-19. In this respect, the pandemic created conditions which Best et al^33^ identify as crucial for successful large-scale change: unified leadership around how to respond, locally driven changes, and rapid monitoring and feedback to enable adaptation in light of emerging problems. Reduction in organisational bureaucracy meant that staff were empowered to identify and rapidly implement changes within and between teams, what May, Johnson and Finch^3^ refer to as conditions of greater organisational plasticity.

This case study demonstrates the productive potential of giving staff agency to develop workarounds for problems based on their local knowledge and experience^43,44^. We join other voices in emphasising the importance of involving those who will deliver change in decisions about how it should be done^28,36^ Rapid change was possible because staff were given the autonomy to decide how to adapt existing working practices. However, we have identified a number of important nuances to this claim.

First, staff were not involved in all decisions about service adaptation. There was minimal consultation of staff involved in service delivery regarding key decisions about cancelling and re-starting elective surgery, despite pre-pandemic policy guidance placing emphasis on hospitals making local decisions about service reorganisation^11–14^. Pressure from COVID-19 was not the same in different Trusts, and staff were frustrated by top-down decisions to cancel non-urgent elective surgery in the absence of critical reflection on alternative service models, as had been done in other countries^21^. Our findings build upon those of a recent scoping review on the immediate and long-term impact of COVID-19 on the delivery of surgical services, which has argued that contingency plans for continuing with surgical care during the pandemic were missing at a global level and this had a negative impact on patient prognoses, outcomes and experience^45^.

Second, a crucial issue was that the solutions created by staff were temporary, emergency responses. Workarounds, such as those employed by participants in this study, are usually time-limited responses to ongoing structural issues^43^.While staff lauded their freedom to innovate and the increased investment of organisational resources, they faced a significant challenge in trying to safely and sustainably design and deliver two parallel systems of care for COVID-19 and non-COVID-19 services. Simultaneously delivering both systems within the resources available to them was felt to be untenable. While staff could draw on their clinical knowledge and experience to adapt services, they had limited practical experience regarding the specific requirements of service delivery under pandemic conditions. There was a perceived absence of organisational knowledge and preparedness in this respect.

Finally, staff were placed in a position where they had to choose which surgeries were urgent and which could wait. These were not simple clinical decisions, but complex ethical choices that had to be contextualised within the lives of their patients^46,47^. The guidance available from national bodies, such as the Royal College of Surgeons^4^, did not sufficiently support staff to engage in these ethical dimensions of care. Struggles to prioritise and cancel surgeries impacted the mental health of HCWs, many who felt overwhelmed and stressed with the backlog of procedures^48^. Furthermore, conflicting or absent guidance from national bodies about safe delivery of surgery during the pandemic led to ongoing anxiety about infection control for themselves and for patients^49^.

### Implications for policy and practice

#### Consistent guidance on pandemic preparedness and response

The inconsistent or contradictory guidance shared with hospital teams created confusion when redesigning service delivery models. Few institutions had organisational knowledge in the form of pandemic preparedness strategies in place. National bodies producing future guidance should ensure that lessons learnt from this and other pandemics are clearly and effectively communicated. This should consider both the ethical and practical dimensions of care-delivery during a pandemic.

#### Staff involvement in decision-making

The absence of staff voices in key strategic decisions about the pandemic response had significant consequences for staff experience and patient care. Previous pandemic response guidance advocated for greater autonomy of decision-making at Trust level. Involving staff in ongoing planning should be a priority, enabling staff to contribute their knowledge and experience to decisions about how to sustainably change service provision.

#### Ensuring staff and patient safety

Policy and guidelines now focus on maintaining non-urgent elective operations through subsequent waves of the pandemic^5^. However, clearing the backlog of surgical procedures is dependent on the ability of staff to safely deliver surgical care. Measures needed to maintain consistent delivery of elective surgical services include safe staffing levels, effective testing of staff and patients and sufficient resources of PPE, drug stocks, theatres and recovery units^50^

#### Maintaining positive innovations

The creative response of staff redesigning services led to a number of valuable innovations, such as improved collaboration within and across surgical units and greater uptake of remote pre- and post-operative care. Ensuring that systems and resources are in place that support the sustainable continued uptake of innovation should be prioritised.

### Study limitations

The findings offered in this paper should be viewed in light the limitations of the study. First, although data were collected during the height of the first wave of the COVID-19 pandemic from a range of staff involved in delivery of surgical services, our respondent pool was mostly comprised of senior staff and was not ethnically diverse. The experiences of more junior staff may have shed light on different issues. Moreover, we only recruited staff from NHS hospitals in London, excluding private hospitals that took on additional NHS surgical work.

Despite these limitations we maintain that the overall lessons taken from the case study provide useful insights into sustainable MSC that can be applied throughout the UK and across other nations with a comparable health care system. Their relevance will be amplified by further research exploring how healthcare service delivery has changed in subsequent waves of the pandemic and in other countries. Moreover, it will be important to examine the sustainability of changes over time to see if those that were identified as positive remain in place.

## Conclusion

In this paper, we argue that both the successes and challenges in the reorganisation of surgical care during the pandemic are related to the involvement of HCWs in decision-making. Our analysis revealed important contradictions in their experiences. On the one hand, staff were empowered to lead decisions about the practicalities of service re-organisation, enabling creative service adaptations in line with local constraints and opportunities. On the other, there was an absence of involvement of staff in wider system-level pandemic decision-making and unclear guidance about how to continue safely delivering surgery and prioritising who needed it. Limited preparedness and lack of staff involvement ultimately led to short-term gains in terms of infection control but a long-term impact on the delivery of certain services Ensuring sustainable MSC requires effective engagement and involvement of those delivering change.

## Data Availability

The authors confirm that the data supporting the findings of this study are available within the article or its supplementary materials.

## Appendix

### Appendix 1 – Topic guide

1. **Background: Can you tell me about your role?**
  - *Can you tell me a bit about your role? (e*.*g. Daily tasks, department, responsibilities)*
2. **Have you been in contact with patients who had suspected and/or confirmed COVID-19?**
  - *In what capacity?*
  - *How have you found working around these patients?*
  - *PPE physical effects? (E*.*g. dehydration, discomfort, restriction in movement, difficulties communicating)*
  - *How has PPE impacted the type of care you provide patients?*
  - *What psychological/emotional impact did this have on you?*
3. **Could you briefly explain how Covid-19 has impacted on elective and emergency surgery at your health facility?**
  - *How has this affected your normal daily tasks/responsibilities? Change of role?*
  - *What tasks are you able to do more or less effectively?*
  - *Has there been an impact on staff’s ability to make diagnoses and act on them?*
  - *Has the supply of drugs, equipment and PPE been affected?*
  - *Have staff been redeployed from or within your health facility*
4. **What do you think about the initial decision to cancel or delay non-emergency surgeries?**
  - *Was it made at the right time?*
  - *How was the decision communicated to you?*
  - *Did you express any concerns about the strategy and if so, were you listened to?*
  - *Have any decisions during Covid-19 have been clinically-led by yourself or colleagues at your facility? Which?/Why not?*
  - *What effect did this decision to delay/cancel surgeries have on care-seeking by patients?*
  - *How do you think the delay or cancellation of surgeries will have impacted on patient outcomes and mental health?*
  - *Have you felt that delaying or cancelling patients’ surgeries has had an effect on your own mental health? In what way?*
5. **What were the preparedness strategies implemented locally (department, hospital or Trust)?**
  - *Did you feel these strategies were enough?*
  - *What do you feel was particularly successful?*
  - *Should the Trust have prepared differently?*
  - *Did you receive any training? (including but not limited to PPE training such as mental health and well-being training)*
  - *Did you have access to guidance on PPE?*
6. **NHS England issued a six-week action plan to restart non-urgent Covid-19 care**.
  - *Are you aware of the procedure for how and when services will restart at your health facility?*
  - *What impact will this have on your role and your daily tasks?*
  - *Are staff being given the opportunity to provide feedback and through what means?*
  - *What do you think about the handling and coordination of this pandemic with regards to surgery? What was done well and what would have been improved?*
7. **Do you currently have any concerns or fears in relation to** …
  - *Restarting elective surgeries (timing, staffing, equipment, PPE, capacity, patient safety)*
  - *Other work concerns (response efforts, PPE, services)*
  - *The national effort*
8. **Over the past months, have you experienced any problems with aspects of your daily life such as sleeping, eating, concentration, or additional worries or anxiety?**
9. **Mental health support (to address risk of moral injury, trauma and developing severe mental health problems)**
  - *Are you aware of any support available for staff wellbeing and mental health?*
  - *Have you had the opportunity to talk about your mental health with your supervisor/team leader?*
  - *Have you had worrying experiences in the last week? Did you receive support after? If so, what type of support? (including formal and informal support)*
  - *Interactions between peers: Do you have time to socialise with your team? What has changed with COVID-19?*
10. **Is there anything else you would like to mention that you feel is important?**

